# SARS-CoV-2 antibody seroprevalence in industry workers in Split-Dalmatia and Šibenik-Knin County, Croatia

**DOI:** 10.1101/2020.05.11.20095158

**Authors:** Ivan Jerković, Toni Ljubić, Željana Bašić, Ivana Kružić, Nenad Kunac, Joško Bezić, Arijana Vuko, Alemka Markotić, Šimun Anđelinović

**Author notes:** These authors contributed equally (Ivan Jerković, Toni Ljubić). Correspondence to: Željana Bašić, Associate Professor, PhD, University Department of Forensic Sciences, University of Split, Ruđera Boškovića 33 21000 Split, Croatia.

## Abstract

**BACKGROUND:** As a result of global spread, COVID-19 has also affected the Republic of Croatia in the last week of February. Although official data show that the number of newly infected is declining, it is still unknown what proportion of the population has been affected by the disease.

**AIM:** To examine seroprevalence of SARS-CoV-2 antibodies in industry workers population sample.

**METHODS:** From 23 to 28 April 2020, we conducted serological testing for antibodies (IgG and IgM) on 1494 factory employees living in the Split-Dalmatia and Šibenik-Knin County (Croatia). We analysed antibody seroprevalence on the level of the company, county, and separately for employees living at the factory premises with limited mobility during the lockdown measures.

**RESULTS:** In a total sample of tested company employees, we detected antibodies in 1.27% of participants (95% CI 0.77-1.98%). In Split facility 13/1316 (0.99%, 95% CI 0.53-1.68%) of participants were tested positive, of which 13/1079 (1.20%, 95% CI 0.64-2.05%) of those living outside the facility and 0/237 (0%, 95% CI 0-1.26%) of those living inside the facility. In Knin facility, 6/178 (3.37%, 95% CI 1.25-7.19%) participants were tested positive for antibodies. The difference between Split (no mobility restrictions) and Knin, was not statistically significant (χ^2^ = 3.47, P = 0.062).

**CONCLUSIONS:** The study showed relatively small SARS-CoV-2 antibody seroprevalence in the DIV Group population sample. When the study findings are interpreted on the county levels, they could indicate that most of the counties’ population was not exposed to the virus.

## INTRODUCTION

Coronavirus disease (COVID-19) is a disease caused by severe acute respiratory syndrome coronavirus 2 (SARS-CoV-2) [1]. Due to its rapid spread across the world, the WHO declared it as a global pandemic on 11 March 2020 [2]. According to the most recent WHO Situation Report on 28 April (when our research was completed), there were 2 954 222 confirmed cases, which led to death in 202 597 cases [3].

On 25 February 2020, the first confirmed COVID-19 case in the Republic of Croatia was reported in a male patient who had recently returned from Italy, which was at that time the major hotspot of the disease in Europe [4-6]. As a response, on 19 March the Croatian Government introduced restrictions that limited social gatherings, operation of shops/services, as well as the prohibition of sporting/cultural events and closing of the borders [7, 8]. Finally, from 23 March, citizens were also prohibited from leaving their place of residence [9]. The response to the COVID-19 crisis by the Croatian Government was seen as one of the most rigorous worldwide, placing it on the top of the stringency scale of the Oxford COVID-19 Government Response Tracker on 26 March [10]. Following the decrease of new daily confirmed cases and decrease of basic reproduction number to 0.8 [11], on 19 April the Croatian Government lifted the restriction of prohibiting citizens from leaving their place of residence. On 27 April, the Croatian Government also started the gradual loosening of measures in an attempt to reduce the negative economic impact [12]. The last official data by the Croatian Institute of Public Health (28 April 2020) reported a total number of 2 055 confirmed cases and 63 deaths [13].

In Split-Dalmatia County (N = 454 798) and Šibenik-Knin County (N = 109 375)[14], the first cases were reported on 15 March [15], and 19 March respectively [16]. According to the data available on the first day of testing (23 April), there were 454 confirmed cases (100 cases per 100 000) in Split-Dalmatia County which made it one of the two most affected counties in Croatia [15], In contrast, on 27 April in Šibenik-Knin County, 83 cases (76 cases per 100 000) were reported [16].

As in all of Croatia, many companies in named counties also had to temporarily reduce or completely stop the production during the restriction measures. However, some of them, employing a great number of people in the county, managed to keep the production in lower quantity by introducing a particular set of protective measures. One of them is the DIV Group, which specialises in the production and trade of screws and other mechanical parts and metal products, as well as shipbuilding [17]. Their two major production sites are located in Split (Split-Dalmatia County), and Knin (Šibenik-Knin County) employing around 2200 people and around 400 people, respectively [17]. The Split facility spreads across around 540 000 m^2^, while the Knin facility comprises the area of about 22 000 m^2^. The employees in both facilities work in different production segments and administration. At the facility in Split, some of the employees live at the facility premises.

Unlike many other businesses, the DIV group introduced protective measures ahead of the Croatian Government. From 25 February, the company implemented hand disinfection stations in all rooms, as well as regular workstation cleaning protocols. All communal coffee and food vending stations were closed as well. From 3 March, all employees had to undergo temperature checks before entering the facility. From 11 March, eight days before national measures took place, they introduced specific internal measures that included self-isolation for those returning from abroad, work from home for part of the business and administrative staff and vacation for employees whose work could not be continued. The total number of employees working at the aforementioned facilities was reduced to around 1300 and 300, respectively. They also continually provided information and warnings for employees using multiple internal communication methods. Before this study, the company facility in Split reported a total of 7 confirmed cases, 20 employees with symptoms of COVID-19, and 52 employees in self-isolation in different periods. However, all the confirmed individuals were in contact with the virus outside off the factory premises while on vacation or working from home.

To preliminarily assess the current state of infection and measures’ efficiency, and to ensure safe conditions, the company management decided to screen their employees for SARS-CoV-2 antibodies. The testing was conducted in Split and Knin company facilities, but also in Zagreb offices (n = 30) and Samobor facility (n = 72), which were not considered in this study due to limited sample size.

Since the prevalence of corona disease in populations is still unknown, it is a priority to gather information from different parts of the world and different target groups. Although RT-PCR tests are reliable to detect current infection and viral material, they cannot provide information on previous infection or exposure to the virus [18]. For this reason, and due to the lower financial and temporal demands, serological immunoassay tests have been recently introduced in studies [19-24]. Previously published studies, as well as unofficial study results, report various proportions of antibodies in studied samples, ranging from 2% to 30% [19-24]. These results could indicate that differences may stem from the sampling strategy as well as the population studied, but also from the performances of serological tests.

The aim of this study was to estimate the proportion of company employees that developed antibodies for the SARS-CoV-2 virus and interpret results to estimate antibody seroprevalence in Split-Dalmatia and Šibenik-Knin County population.

## METHODS

We conducted serological immunoassay testing for SARS-CoV-2 antibodies in 1494 adult employees of DIV Group, in Split 1316 and Knin 178. The study was performed between the23 to 28 of April 2020, in the last days before the loosening of national restrictive measures in Croatia. DIV Group organised the study in cooperation with the Clinical Department for Pathology, Forensic Medicine and Cytology, University Hospital Center Split; University Department of Forensic Sciences, University of Split; and University of Split School of Medicine (Split, Croatia).

To examine whether the results could be generalised to the counties’ populations, the required sample size was calculated using the online sample size calculator (https://www.surveymonkey.com/mp/sample-size-calculator/) with a confidence interval of 95% and the margin of error of 5%. For Split-Dalmatia County (N = 454 798) the required sample size was 384, while for the Šibenik-Knin County (N = 109 375), it was 383.

### Participants and recruitment

DIV Group management and its crisis headquarters invited employees to participate in voluntary screening for SARS-CoV-2 antibodies via phone and/or e-mail. The schedule for the screening was arranged in cooperation with the head of company departments and sub-companies. Each participant was provided with a questionnaire that had to be completed prior to the test and was asked to sign an informed consent form. The questionnaire form contained questions on basic demographic data, symptoms, recent travels abroad, and possible contacts with infected or likely infected individuals.

### Test type and performance

We used AMP Rapid Test SARS-CoV-2 IgG/IgM (AMP Diagnostics, AMEDA Laboradiagnostik GmbH, Graz, Austria). The test is intended for the rapid immunochromatographic qualitative detection of IgG and IgM antibodies to SARS-CoV-2 in whole human blood, serum, and plasma samples. According to the manufacturer, for IgM, test sensitivity is 95.7%, test specificity 97.3%, and test accuracy 96.8%. For IgG, test sensitivity is 91.8%, test specificity 96.4%, and test accuracy 95%. Information on the combined test performance (IgM and IgG) was not provided by the manufacturer [25].

The previous study [26] addressed that even when test performance parameters are high, test results can be largely affected if incidence level in population is low. For example, the authors showed that an assumed test, with a sensitivity of 75% and specificity of 95%, could provide 640 positive and 9360 negative cases on a population of 10 000 with an incidence base rate of 2%. Out of the 640 positive cases, due to the test performance, 150 cases would be truly positive, while the other 490 would be truly negative. In contrast, out of the 9360 negative cases, 9310 would be truly negative, while 50 would be truly positive. The probability of a truly positive case yielding a positive test result would be calculated as the ratio between truly positive cases and all positive test results. A similar approach could be used to calculate the probability of a truly positive case that showed negative test results, as a ratio of truly positive cases tested negative and all cases with negative test results [26]. In this hypothetical case, the probability of a truly positive case testing positive would be 150/640, which would mean that only 23.44% of participants tested positive actually are positive cases. In contrast, the probability of truly positive cases testing negative would be 50/9360, which would imply that 0.53% of the participants tested negative would actually be positive [26].

For this reason, before conducting the study, we estimated the probabilities that a participant really has antibodies if tested positive or negative. For this calculation, we used the test performance data specified by the manufacturer and assumed that in the sample of 2000 people, that was the expected size of the study sample, one per cent of participants would have IgG, and one per cent of participants would have IgM antibodies against SARS-CoV-2.

With the parameters stated above, we estimated that 20.22% of the sample tested positive for IgG could really have IgG antibodies, while 0.1% of the sample tested negative would have IgG antibodies. IgM antibodies could be really present in 26.39% of the sample tested positive for IgM, and 0.05% of the sample tested negative.

### Ethical approval

Ethical approval was attained by the University Department of Forensic Sciences Ethics Committee on 22 April 2020 (2181-227-05-12-19-0003; 024-04/19-03/00007).

### Sample collection and testing

In Split, the screening tests were performed inside the company classroom (about 80 m^2^), with four testing points arranged in semi-circular pattern placed two or more meters away from each other. In Knin, two testing points were established in a company classroom (about 60 m^2^) at a distance larger than 4 meters. Both rooms had a separate entrance and exit points and were manned by a security guard, that was in charge of escorting the participants into the room and checking whether they had protective equipment (face mask). During the screening, the rooms were continuously ventilated.

For each participant, we punctured a finger with a sterile lancet and collected approximately 10 μL of whole blood with a disposable pipette. The blood was immediately transferred to the sample well of the test cassette, after which approximately 80 μL of buffer was applied in the buffer well. According to the manufacturer’s recommendations, the test results were (C - control line; G - test line IgG; M - test line IgM) read between 10 and 15 minutes. Each invalid test (no reaction on control line) and each positive test result was repeated at a different testing point (different test performer, lot, and buffer). All test results were entered into the form and photographically documented in the recommended time interval. If a participant tested positive, they were forwarded to the local epidemiologic service, where they would undergo additional confirmatory testing using RT-PCR. Crisis headquarters maintained further contact with physicians and epidemiologic services to receive information on final results. Additional information about the whereabouts and possible contacts of the positive participants were provided to the researchers by the crisis headquarters.

### Statistical analysis

We provided descriptive statistics on population demographic structure. We calculated the raw proportion of positive test results in the population sample as well as a proportion on a 95% confidence interval. To calculate two-sided 95% CI, Clopper-Pearson exact method in RStudio (version 1.2.5033, RStudio, Inc., Boston, MA, USA) and package GenBinomApps (https://CRAN.R-project.org/package=GenBinomApps) were used. We analysed the results in the complete sample and separately according to two counties. In the facility in Split, we also analysed separately the participants who lived on the facility’s premises and were limited to leave the facility more than once a week during the national restrictive measures, and the participants living outside the facility. Differences between prevalence in different counties were analysed using a chi-squared test with Yates correction (due to the low number of expected frequencies). The level of statistical significance was set at P ≤ 0.05.

### Funding

The tests used in this study were purchased by the DIV Group.

## RESULTS

### Participants’ characteristics

The tested population comprised a wide age population structure, ranging from young adults to elderly population, mostly working in the company as senior consultants and engineers. In total, 1494 participants (88.1% male; median age 46, range 18-79) from the DIV group were tested. In Split, there were 1316 participants (89.4% male; median age 46, range 19-79), while in the Knin facility, there were 178 participants (78.1% male; median age 45, range 18-64). Of the total number of the participants in Split, 237 (18%) were those accommodated inside the facility grounds.

### Testing performance

A total of 1521 immunoassays were used in the study. Six of them were repeated as they did not show a reaction in the control region, while 21 tests were repeated for participants who showed positive first test results. When repeated, all tests showed a reaction in the control region, and tests that were repeated for positive participants demonstrated the same results.

### Positive test results

Table 1 shows an overview of the screening test results and sample sizes, divided by counties and degree of mobility restrictions.

**Table 1.**
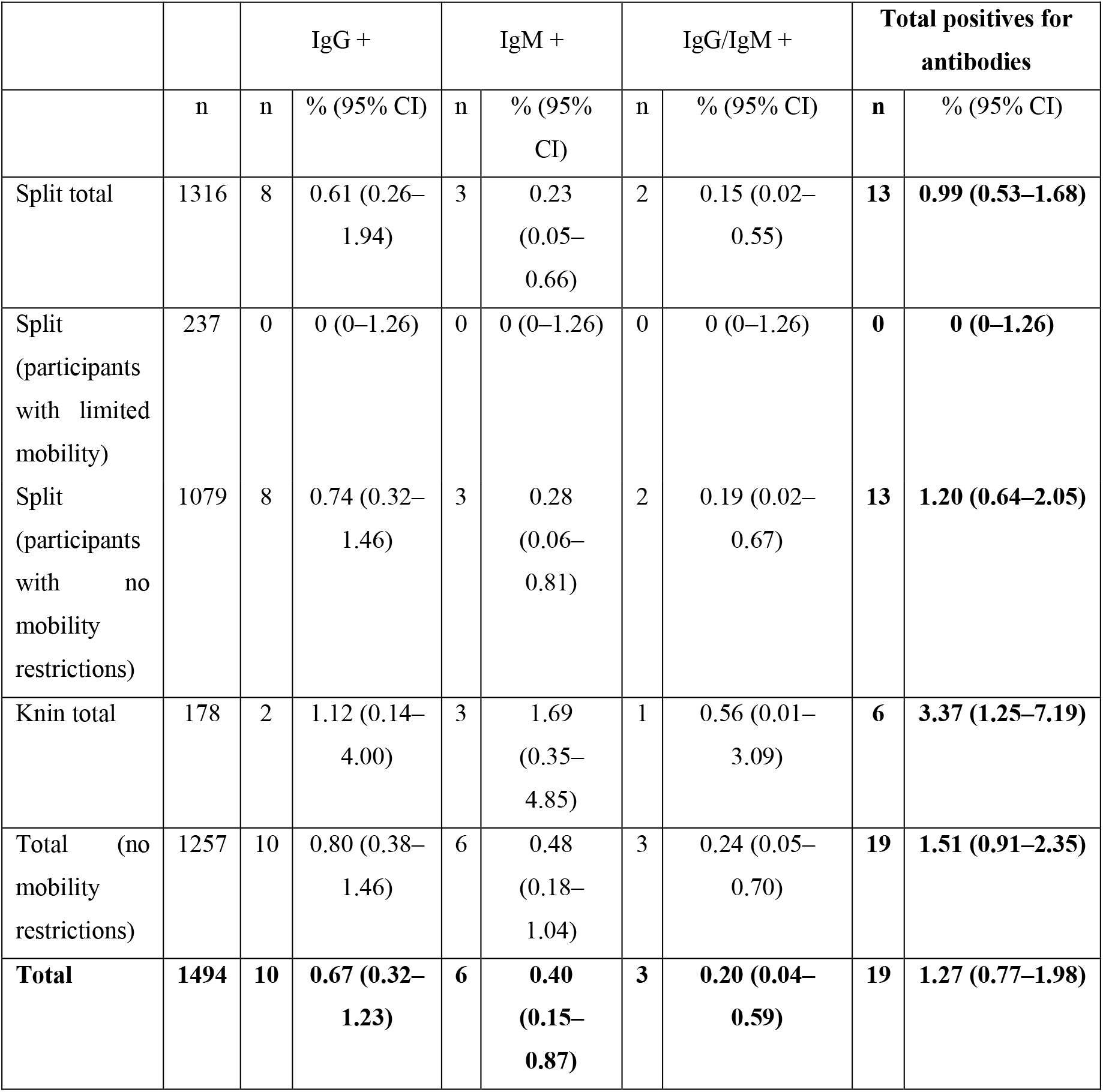
SARS-CoV-2 antibody prevalence according to the analysed population samples

The seroprevalence of SARS-Cov-2 antibodies in the tested population sample was 1.27%, but ranged from 0 to 3.37%, depending on which population subsample was considered. In the total population sample, the proportion of positive individuals was highest for IgG, followed by IgM and a combination IgG/IgM antibodies, respectively. Positive test results were detected for 17/1316 males (12 from Split and five from Knin facility) and 2/178 females (one from Split and one from Knin facility). Both females were positive only for IgM antibodies.

All participants living inside facility premises, and with limited mobility during the lockdown measures, tested negative for antibodies. When we excluded them from the overall sample, the proportion of positive participants slightly increased and reached 1.51%.

In the Split facility (Split-Dalmatia county), antibodies were detected in 0.99% of the sample, for the total number of employees, and 1.20% for the employees living outside the facility, whereas in Knin (Šibenik-Knin county) seroprevalence of antibodies was 3.37%. Difference between proportions of positive participants in Split (with no mobility restrictions) and Knin samples, was not statistically significant (x^2^ = 3.47, P = 0.062).

### Positive participants

From all positive participants, three of them (one IgM, two IgG/IgM) reported close contact with a person suspected of SARS-CoV-2 infections or with a confirmed case. Table 2 shows symptoms reported to occur since the beginning of 2020 by positive participants.

**Table 2.** Sex, age, and symptoms reported by positive participants

| Sex | Age | Antibodies | Symptoms |  |  |  |  |  |  |
| --- | --- | --- | --- | --- | --- | --- | --- | --- | --- |
|  |  |  | General weakness | Nose leak | Sore throat | Cough | Headache | High body temperature | Muscle pain |
| Split |  |  |  |  |  |  |  |  |  |
| M | 43 | IgM | - | - | - | + | - | - | - |
| M | 59 | IgG | - | - | - | - | - | - | - |
| M | 59 | IgG | - | - | - | - | - | - | - |
| M | 54 | IgG/IgM | - | - | - | - | - | - | - |
| M | 59 | IgG | - | - | - | - | - | - | - |
| M | 46 | IgG | - | - | - | - | - | - | - |
| F | 26 | IgM | + | - | - | - | + | - | - |
| M | 49 | IgG | - | - | - | - | - | + | - |
| M | 64 | IgG | - | + | - | + | - | - | - |
| M | 55 | IgG/IgM | - | + | - | - | - | - | - |
| M | 42 | IgG | - | - | - | - | - | + | + |
| M | 51 | IgG | - | - | - | - | - | - | - |
| M | 53 | IgM | - | - | - | - | - | - | - |
| Knin |  |  |  |  |  |  |  |  |  |
| M | 60 | IgG | + | + | + | - | + | + | + |
| M | 25 | IgM | - | - | - | - | + | - | - |
| M | 62 | IgG/IgM | - | - | - | - | - | - | - |
| M | 52 | IgG | + | + | - | - | - | - | - |
| M | 46 | IgM | - | - | - | - | - | - | - |
| F | 56 | IgM | - | + | + | - | - | - | - |
| Symptoms total: |  |  | 3 | 5 | 2 | 2 | 3 | 3 | 2 |

All IgM positive participants were proceeded to confirmatory testing (RT-PCR). According to feedback received, from a total of six participants with IgM antibodies, two of them were positive for SARS-CoV-2 on RT-PCR, while four participants tested negative.

## DISCUSSION

The present study showed that seroprevalence of SARS-Cov-2 antibodies in the tested company population sample, ranges with different inclusion criteria, from 0.77 to 1.98% and from 0.91 to 2.35%. Due to the sample size, these results could also reflect seroprevalence in the general Split-Dalmatia County population, thus indicating the relatively small proportion of population exposure to the virus. To the authors’ knowledge, this is one of the few reported studies of seroprevalence of SARS-CoV-2 antibodies using immunoassay tests, that have only been introduced recently [20-22].

The present research was primarily aimed at the population working at DIV Group facilities in Split and Knin and showed that 0.77 to 1.98% of the population had developed antibodies. As it was previously mentioned, some of the participants from Split are living inside the facility with more restrictions. Although differences between the Split sample living inside and outside the facilities could not be statistically compared, the results could suggest that the restriction of movement outside the facility (complete lockdown) additionally lowered the transmission factor.

Overall, results on the company population level could be attributed to the early implementation of company protective measures combined with strict national measures, put in place to mitigate the COVID-19 spread.

Although the research was conducted on the seemingly closed population samples, the majority of the participants, excluding those accommodated on the facility grounds, could in part represent a general county population or population that is slightly more exposed to the virus due to the greater interpersonal contact outside their homes (as they continued to work during the shutdown). In this sample, which considered both counties together, the seroprevalence for antibodies was 1.51% (95% CI 0.91-2.35%).

In the sample from Split-Dalmatia county, which was one of the two most affected counties in Croatia (measured in virus incidence per 100 000), the study revealed that antibodies were present in 1.20% (95% CI 0.64-2.05%) of the population living outside the facility grounds. Even though the sample was not stratified, it comprised a representative sample size for the Split-Dalmatia County population, which could reflect a relatively realistic antibody seroprevalence in the county.

The study also provided results for Šibenik-Knin County, which could imply a higher, but not statistically significant, prevalence of the antibodies (CI 95%, 1.25-7.19%). Due to the restricted sample size, that does not meet the criteria to be a representative sample for Šibenik-Knin county, these results should be treated only as preliminary findings.

Due to the limited sample size from the Šibenik-Knin County, we decided to consider only results from Split-Dalmatia county, excluding the participants living on the facility grounds. When only this population sample is considered, the antibody seroprevalence was 1.20% (95% CI 0.64-2.05%).

Until now, only a few serological study results have been reported, but due to different sampling strategies and population, they are not directly comparable to our study. For example, a study conducted in Santa Clara County (N = 1 928 000), California, USA [20], which aimed to target the general population, found antibodies in 1.5% (95% CI 1.11-1.97%) of the population sample. A study conducted on a general population sample of the island of Jersey, UK (N = 106 800), included a total of 855 participants from 438 households. The results showed that the seroprevalence of SARS-CoV-2 antibodies is 3.1% (95% CI, 1.8-4.4%) [24]. One of the studies, conducted in Gangelt county (N = 12 529), which was a viral hotspot in Germany, comprised a sample of 1000 participants from 400 households. The results stated that 15% of the population developed antibodies to the virus [21]. One study was conducted in Oise in France that targeted participants from virus affected high school (pupils, their family members, and school staff). In the mentioned study, the seroprevalence of antibodies was 25.9% (95% CI 22.6-29.4) [22].

Also not directly comparable, due to the application of SARS-CoV-2 sequencing instead of serological testing, a study conducted on the Icelandic population sample provided comprehensive results. It was conducted on approximately 6% of the total Icelandic population and showed different results depending on the sampling patterns [27]. For the targeted population with symptoms or high risk of infection, this study found 13.3% positive cases, while open invitation and random sample provided 0.8% (95% CI 0.6-1) and 0.6% (95% CI 0.3-0.9) of positive cases, respectively.

The major limitation of the study was the characteristics of serological immunoassay tests, which are currently not sufficiently explored and validated [18]. In our case, the most pronounced drawback of the test reflected in the fact that RT-PCR confirmed two of six IgM positive cases (33%), which was not reported in previous serological studies [20-22]. However, due to the generally low incidence of the disease, this result was not unexpected [26]. Specifically, as previously mentioned in our estimation (see Methods section), we could expect that 26.39% of individuals with positive test results would truly have IgM antibodies. However, it does not imply that people tested negative truly have IgM antibodies, as our previous calculations estimated that it would be the case only in 0.05%, i.e., in no more than one individual within our overall sample.

To further explain test inconsistencies, other factors should also be considered. Firstly, most of the tests are still not validated, and, as in our case, only manufacturer data about test performance is available. Along with test performance indicators, test manufacturer also stressed that samples with higher heterophile antibodies or rheumatoid factor could affect the test result [25]. Secondly, WHO stated that serological tests, in general, could be susceptible to cross-reaction with other frequent infections, like human coronaviruses causing common cold [28]. Nonetheless, despite their limitations, these tests can still be a valuable qualitative research tool [18]. Due to their low probability rates of positive cases that tested negative [26], they could provide credible information about the proportion of the population that was not exposed to the virus [18].

In conclusion, our study indicated that the proportion of the Split-Dalmatia population that did not develop antibodies to the SARS-CoV-2 virus could be relatively high.

Restrictions that the DIV group implemented, along with national restrictive measurements, had an enormous impact on the suppression of virus spreading. Interestingly, the workers that did not leave the facility during the pandemic did not exhibit any case of the infection. It could be an additional proof that the quarantine is extremely effective, in the same manner, it was introduced in 1377 in Dubrovnik, as the first documented case of quarantine in the history [29]. Thus, Croatian heritage in a way once again contributes to the civilisation.

## Data Availability

The data that support the findings of this study are available from the corresponding author, [ŽB], upon reasonable request.

## Acknowledgements

The authors wish to thank DIV Group company and Tomislav Debeljak, along with all study participants. We are particularly thankful to Boško Ramljak, Marija Čečuk, Ivica Sinovčić, Maja Matić, Milena Matulić, Dražen Bulić, Ružica Jerković, Andrea Kolić, and Rino Rivi Kolombatović for assistance with study organisation and data collection. We are grateful to Prof. Ana Marušić for critical reading of the manuscript. We also thank Marino Krstulović for valuable advice about the manuscript style and proofreading.

## Authors contributions

All authors conceived and designed the study; IJ and TLJ acquired the data; all authors analysed and interpreted the data; IJ, IK, TLJ, and ŽB drafted the manuscript; all authors critically revised the manuscript for important intellectual content; all authors approved the version to be submitted; all authors agree to be accountable for all aspects of the work.

